# Assessing the Mortality Impact of the COVID-19 Pandemic in Florida State Prisons

**DOI:** 10.1101/2021.04.14.21255512

**Authors:** Neal Marcos Marquez, Aaron Littman, Victoria Rossi, Michael Everett, Erika Tyagi, Hope Johnson, Sharon Dolovich

## Abstract

**Background:** The increased risk of COVID-19 infection among incarcerated individuals due to environmental hazards is well known and recent studies have highlighted the higher rates of infection and mortality prisoners in the United States face due to COVID-19. However, the impact of COVID-19 on all-cause mortality rates in incarcerated populations has not been studied.

**Methods:** Using data reported by the Florida Department of Corrections on prison populations and mortality events we conducted a retrospective cohort study of all individuals incarcerated in Florida state prisons between 2015 and 2020. We calculated excess deaths by estimating age-specific expected deaths from mortality trends in 2015 through 2019 and taking the difference between observed and expected deaths during the pandemic period. We calculated life table measures using standard demographic techniques and assessed significant yearly changes using bootstrapping.

**Findings:** The Florida Department of Corrections reported 510 total deaths from March 1, 2020 to December 31, 2020 among the state prison population. This was 42% higher (rate ratio 1.42, 95% CI 1.15 to 1.89) than the expected number of deaths in light of mortality rates for previous years. Reported COVID-19 deaths in a month were positively correlated with estimated excess deaths (80.4%, p <.01). Using age-specific mortality estimates, we found that life expectancy at age 20 declined by 4 years (95% CI 2.06-6.57) between 2019 and 2020 for the Florida prison population.

**Interpretation:** The Florida prison population saw a significant increase in all-cause mortality during the COVID-19 pandemic period, leading to a decrease in life expectancy of more than four years. Life years lost by the Florida prison population were likely far greater than those lost by the general United States population, as reported by other studies. This difference in years lost highlights the need for increased interventions to protect vulnerable incarcerated populations during pandemics.

**Funding:** Vital Projects Fund, Arnold Ventures, US Centers for Disease Control, Eunice Kennedy Shriver National Institute of Child Health and Human Development

## Introduction

The heightened risk of COVID-19 infection and related mortality among prison populations is well documented.^1–5^ Previous studies have estimated that people in prison across the United States are 5.5 times more likely to be infected with COVID-19, and that their age-standardized COVID-19 mortality rate is 3.0 times higher than that of the general population.^1^ While these differences are striking, analyzing COVID-19 infections and reported COVID-related mortality events alone may not fully capture the health burden experienced by prison populations, as it does not analyze the potential impact on total mortality.

Numerous studies have found that, in the general United States population, all-cause mortality has been significantly elevated during the COVID-19 pandemic, providing evidence that COVID-19 deaths are not simply substituting for other deaths that were likely to occur under typical circumstances.^6,7^ For example, a recent study found that, in the state of Florida as a whole, mortality rates were 15.5% higher than expected in 2020 in light of observed mortality trends.^8^ As of yet, however, no research has evaluated whether prison populations have experienced similarly heightened all-cause mortality rates.

We know that crude mortality rates in prisons increased between 2001 and 2016, although this change appears to be largely attributable to an aging prison population, given that age-specific mortality rates declined during this period.^9^ Among individuals 45 and older, leading causes of death in prison include cardiovascular and respiratory related causes,^10^ with prevalence of associated diseases exceeding that in the U.S. population more generally.^11,12^ Although cardiovascular and respiratory diseases are known to lead to increased risk of COVID-19-related morbidity and mortality,^13–15^ it is uncertain whether, given the heightened prevalence of these conditions in the prison environment, deaths in prison from non-COVID-19 related causes during the pandemic would decrease (due to competing risks), increase (due to added risk associated with COVID-19), or remain similar. To date, studies quantifying how the overall mortality rate of incarcerated populations has changed during the COVID-19 pandemic are extremely limited.^16^

Studying mortality events in incarcerated populations in the U.S. is difficult because of limited availability of mortality and demographic information. Delays in the reporting of deaths in prisons make timely evaluations of mortality rates difficult and, to date, comprehensive data on deaths in U.S. prisons are reported only for years up to 2016, with little demographic detail.^17^ An added complication is that data on prison population demographics are sparse, making it difficult to construct appropriate demographic denominators to evaluate mortality rates. This challenge is especially consequential during the COVID-19 pandemic, when prison systems saw striking declines in total population. A recent study found that, between June 2019 and June 2020, prisons across the United States saw a nearly 10% decrease in total population.^18^ Given the politics surrounding prison releases, population declines are likely to be differentially distributed across age and sex. However, demographic compositional shifts in prisons during this time have not yet been studied; as a consequence, age-specific mortality denominators among prison populations are not up to date.

To overcome this knowledge gap, we leveraged access to unique datasets about the Florida prison population to better understand the impact of the COVID-19 pandemic on all-cause mortality. We focused on the state of Florida for two reasons. First, the Florida state prison system was the third-largest state prison system in the United States as of late 2020.^18^ With over 80,000 people incarcerated in Florida prisons at a given time, the population is sufficiently large to understand with some precision the historical dynamics of mortality rates and how they may have been impacted by the COVID-19 pandemic. Second, assessment of all-cause mortality rates in a quickly changing population--as exists in U.S. prisons--requires temporally detailed data on mortality events and population structure. Florida is one of the few state prison systems for which available data allow us to evaluate mortality rates in the pandemic period and compare these rates to those observed in previous years.

Our main goals for this analysis were to 1) document demographic changes that occurred in the Florida state prison population during the COVID-19 pandemic, 2) assess excess deaths that occurred during the months of the COVID-19 pandemic in 2020 and their temporal relationship to reported COVID-19 deaths, and 3) quantify changes in the overall mortality rate of the Florida state prison population in 2020.

## Methods

### Study Population

As this study did not meet the definition of Human Subjects research and all data is publicly available, IRB approval was not sought. The population analyzed for this study consists of those individuals incarcerated from 2015 through 2020 in Florida state prisons. The Florida state prison system, administered by the Florida Department of Corrections (FLDOC), comprises over 50 main institutions with a current aggregate daily population of over 80,000 individuals.^18^ While FLDOC has produced mid-year population reports through 2019 with varying levels of demographic detail, those reports provide only a snapshot of the population on a single day and do not capture the changes in the population which occur over the course of the year. Proper assessment of the person-time risk of mortality requires more temporally-detailed demographic data.

To construct precise, time-specific demographic profiles of the Florida prison population, we reconstructed monthly population counts back to January 2015 using publicly available FLDOC inmate population records. Because the intended use of the inmate population records is not to create a point-in-time population estimate, we compared our estimates of the population to the figures reported by FLDOC in its yearly mid-year report by age-groups and sex. We found that our total population estimates for those dates did not differ from reported figures by more than 1.5%, while our estimates of the percentage of the population share by age-groups and sex do not differ by more than 1 percentage point.

Our population dataset consists of age- and sex-specific monthly population data for all individuals in the Florida state prison system age 20 and above between January 2015 and December 2020. In total, we analyzed 6,830,581 person-months of data. Yearly breakdowns of population counts may be found in Table 1.

**Table 1:** Descriptive statistics of Florida state prison population by year

| Year | Person-Months | Median Age<br>Dec 31 | % Female<br>Dec 31 | Deaths | Death Rate<br>Per 100K<br>Person-Years |
| --- | --- | --- | --- | --- | --- |
| 2015 | 1,163,659 | 36.9 | 7.24 | 353 | 364 |
| 2016 | 1,154,419 | 37.3 | 7.18 | 356 | 370 |
| 2017 | 1,143,776 | 37.7 | 7.12 | 428 | 449 |
| 2018 | 1,139,271 | 38.2 | 7.24 | 440 | 463 |
| 2019 | 1,146,362 | 39.0 | 7.23 | 400 | 419 |
| 2020 | 1,083,104 | 40.2 | 6.66 | 590 | 654 |

### Mortality Reporting

All-cause mortality of the Florida state prison population is reported by FLDOC. FLDOC publicly releases a range of information concerning all mortality events in the Florida state prison population, including decedents’ sex, date of birth, and date of death. Information on the manner of death is also sometimes provided; however, these indicators are not standardized to uniform causes of death (such as those provided by the International Classification of Diseases), so they are not suitable for use in statistical analysis. FLDOC updates these data retrospectively and, in order to avoid undercounting of deaths due to reporting delay, we use death data only for dates through the end of 2020.

### COVID-19 Deaths

Although FLDOC reports the cumulative number of deaths attributable to COVID-19 within the state prison population to date, it does not report historical trends or monthly breakdowns pinpointing when these deaths occurred. To create such a time series, we leveraged data we collected at the UCLA Law COVID-19 Behind Bars Data Project, where we catalog and publish officially reported cumulative COVID-19-related infections and deaths in prison populations across the United States. We calculated monthly COVID-19 related deaths back to the beginning of the pandemic by taking the difference in cumulative counts between the last observed value of each month. FLDOC does not release demographic data about the individuals who died from COVID-19.

### Analysis

Annual crude mortality rates were calculated by dividing the number of deaths by person-years observed in a given year. To assess the need to adjust for changing population structure, we calculated the proportion of the population that was female and the median age of the population in each month. Across all months, the proportion of the population that was female stayed within a range of 6.6% to 7.4%. Because of this minimal fluctuation (<1%), we did not adjust in our analyses for changes in the sex composition of the population over time. However, the median age of the population increased substantially and consistently over time, moving from 36.9 to 40.2 years. Yearly data summaries of the Florida prison population are presented in Table 1 and age-specific changes between 2019 and 2020 are presented in Table 2.

**Table 2:** Change in population and mortality rate by age

| Age Group | Population<br>Dec 1, 2019 | Population<br>Dec 1, 2019 | Population %<br>Change | Mortality Rate<br>Ratio '20/ '19 |
| --- | --- | --- | --- | --- |
| 20-24 | 6,931 | 5,011 | -27.7% | .631<br>(CI 0.00 -- 1.96) |
| 25-34 | 28,958 | 23,438 | -19.1% | 1.08<br>(CI .593 -- 1.80) |
| 35-44 | 26,257 | 23,431 | -10.8% | 1.77**<br>(CI 1.13 -- 2.72) |
| 45-54 | 17,870 | 16,323 | -8.6% | 1.05<br>(CI .733 -- 1.48) |
| 55-64 | 11,308 | 11,140 | -1.49% | 1.49**<br>(CI 1.13 -- 1.92) |
| 65-74 | 3,495 | 3,575 | 2.29% | 1.65**<br>(CI 1.28 -- 2.16) |
| 75+ | 717 | 737 | 2.79% | 1.61**<br>(CI 1.18 -- 2.23) |

To account for the changing age structure of the population, we divided our analysis into age groups similar to those used in the Florida Department of Health’s vital statistics reporting.^19^ We omitted people below age 20 due to their small numbers and limited number of deaths (<5) in Florida state prisons from 2015 through 2020. We combined the age groups of 75-84 and 85+, in which mortality rates were high, because of the relatively small population in the latter group.

To assess excess mortality in the pandemic period and the relationship to COVID-19 deaths, we used age-specific monthly mortality data from 2015-2019 to build a baseline measure of the expected mortality rate. We modeled our process on a recent analysis of excess mortality for the Florida population more generally.^8^ We first examined the monthly crude mortality rate using auto-correlation functions to evaluate the need to account for either monthly or seasonal auto-correlation. We found significant evidence of monthly auto-correlation but no evidence of seasonal auto-correlation. We modeled age-specific monthly mortality probabilities independently for each age group using Bayesian hierarchical logistic regression models where, for each month, persons present at the beginning of the month represent the number of trials, and deaths occurring among those individuals represent events. We adjust for monthly auto-correlation in the observed mortality rates by including a latent variable following a random walk structure of order 1. In addition, we include an unstructured independent and identically distributed normal latent variable to account for unstructured overdispersion of monthly probabilities. A full specification of the model including hyper-prior specification is included in the appendix.

Expected mortality for each month was calculated by taking predicted probabilities of mortality for each month and age group in 2020 from the aforementioned models, multiplying the probabilities by the observed monthly populations for the age groups, and summing across all age groups. Excess mortality in the pandemic period was then calculated by taking the difference between observed and predicted deaths for each pandemic month (March 2020 and thereafter) and aggregating across all months to get total excess deaths during the COVID-19 pandemic period. The excess death rate is then simply the excess deaths divided by the expected deaths. This process was repeated for the median, 97.5 quantile, and 2.5 quantile of the posterior of predicted probabilities to generate uncertainty measures. To assess the accuracy of our expected mortality forecast, we evaluate this model-building procedure using data from 2015 to 2018 to calculate excess deaths in 2019, given that it is a year not expected to have a significant percentage of excess deaths.

We calculated the correlation of reported monthly COVID-19 deaths with monthly median estimates of excess deaths in Florida state prisons and evaluated the significance of the correlation using a simple linear bivariate regression test. Estimates of monthly excess mortality and reported COVID-19 deaths are shown in Figure 2.

**Figure 1:**
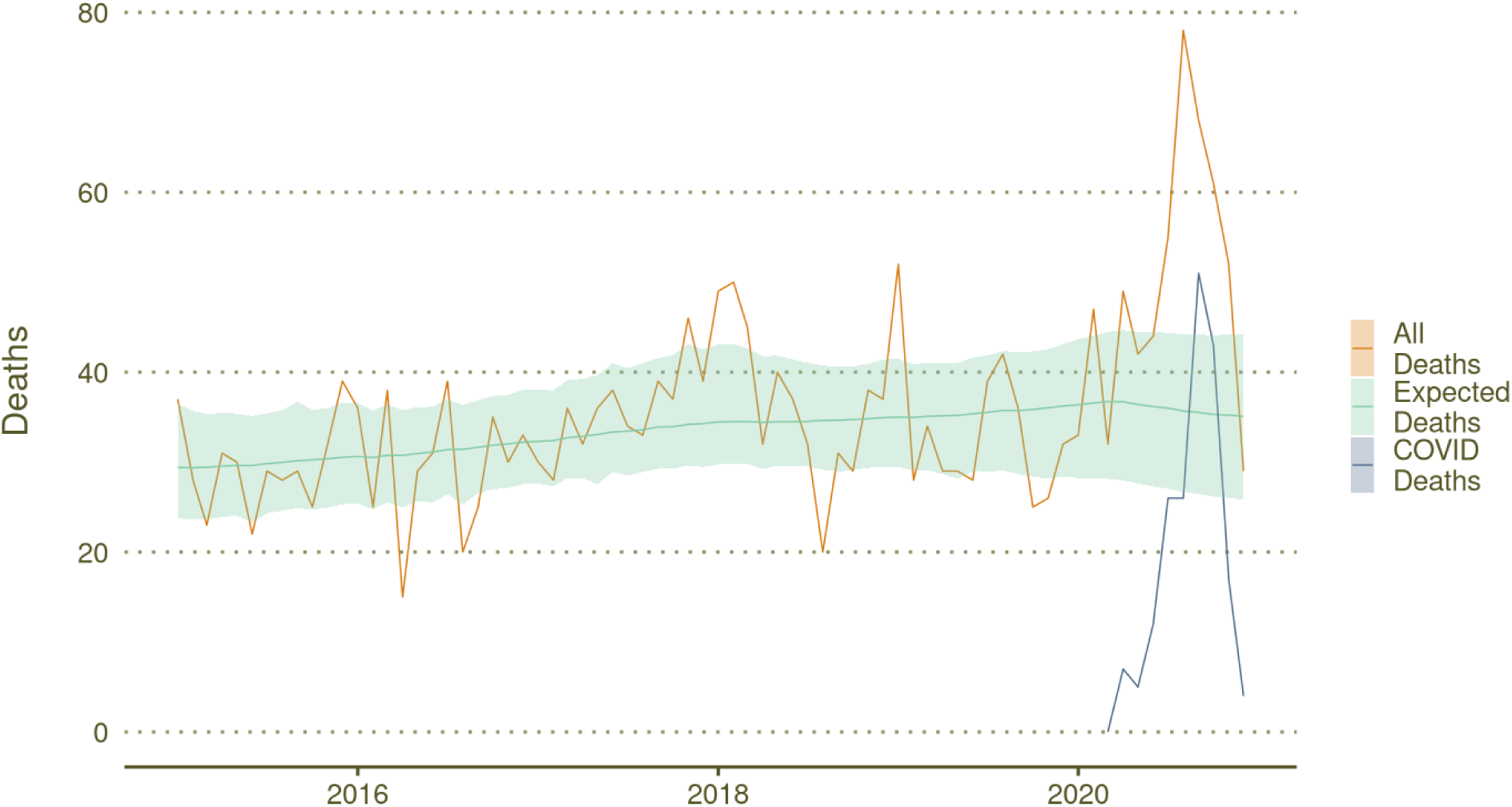
Observed and expected mortality trends in Florida state prison population

**Figure 2:**
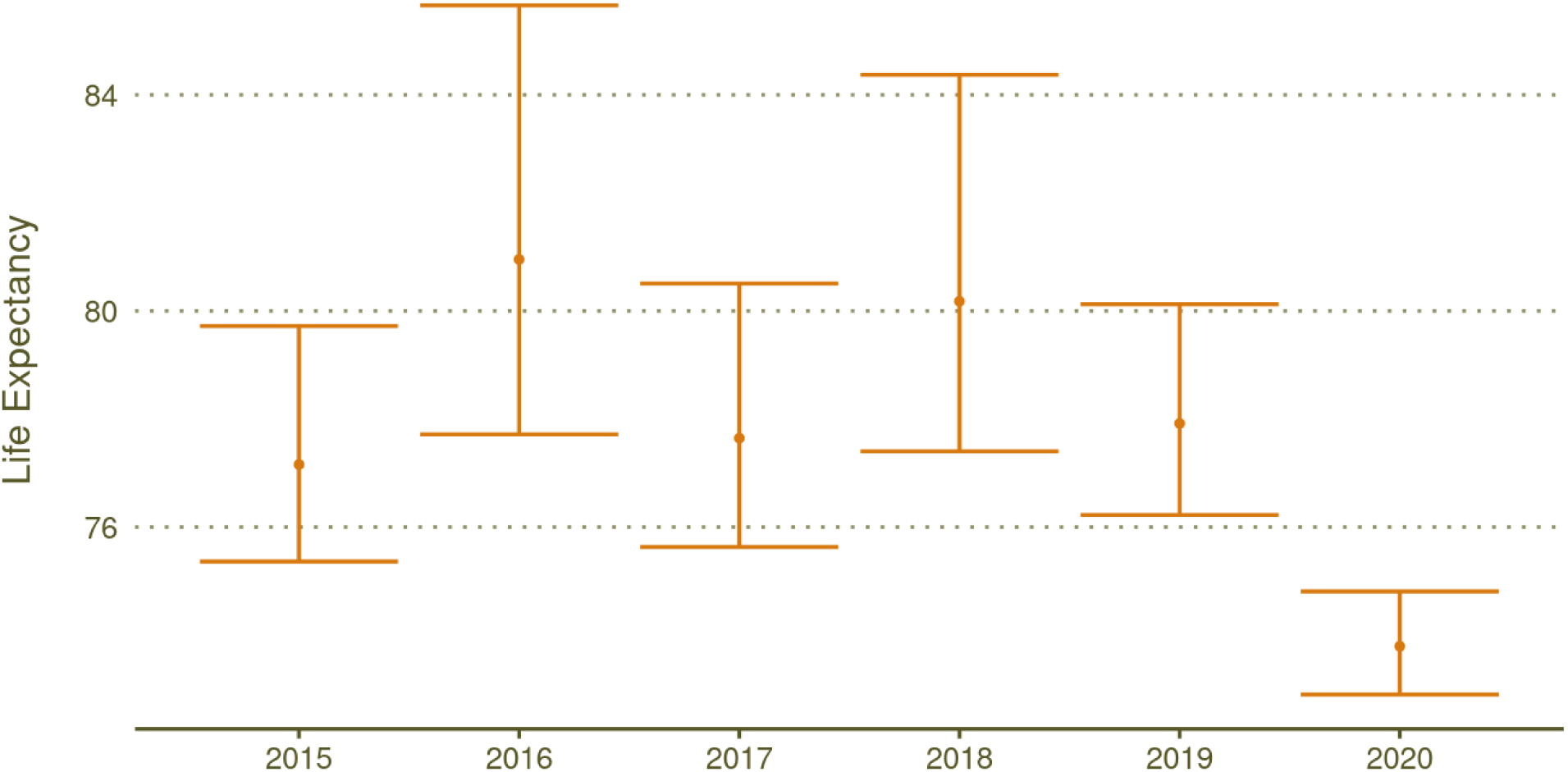
Bootstrapped estimates of life expectancy for Florida state prison population by year

Life table values for each year of analysis were calculated using life table methods outlined in previous demographic work.^20^ Given the low number of deaths that occur in some age groups, we used bootstrapping to obtain measures of uncertainty for life table metrics, taking 10,000 bootstrap samples. Utilizing bootstrapping allows a better understanding of whether the differences in yearly observed calculations of life table measures are simply due to random chance or whether they represent a meaningful change. Our primary measures of interest are age-specific estimates of mortality rate and period life expectancy at age 20—henceforth referred to as life expectancy—as well as the annual change in these values.

Bayesian hierarchical models were fitted with Laplace approximation of posteriors using the INLA package 20.03.17 and all analyses were done in R version 4.04.

## Results

During the study period (i.e., 2015-2020), we observed a total of 2,567 deaths among the Florida state prison population. Comparing crude mortality rates, we found that 2020 saw the highest rate of mortality across all years, with 654 deaths per 100,000 person-years. Furthermore, crude mortality for 2020 was significantly higher than 2019’s crude mortality rate when using bootstrapping to compare crude rates (RR 1.56, 95% CI 1.39-1.76).

Our model for excess deaths found that using data from 2015-2018, there was—consistent with expectations—not a significant presence of excess deaths in 2019. When repeating this process for training years of 2015-2019 and evaluating only the 10 months of the COVID-19 pandemic in 2020, we found that there was a significant number of excess deaths, totaling 42% of expected deaths (95% CI 15.0% - 89.4%). Monthly median posterior estimates of excess mortality were found to be strongly and significantly correlated with monthly reported deaths related to COVID-19 (80.4%, *p* <.01).

Age-specific mortality rate estimates calculated from life tables were found to be significantly higher for four age groups between 2019 and 2020. The age groups of 35-44, 55-64, 65-74, and 75+ saw significant increases in their mortality rates year over year, with rate increases of 77% (RR 1.77, 95% CI 1.13 to 2.72), 49% (RR 1.49, 95% CI 1.13 to 1.92), 67% (RR 1.65, 95% CI 1.28 to 2.16), and 61% (RR 1.61, 95% CI 1.18 to 2.23), respectively. The age group of 45-54 also saw a year-to-year increase, but the change was not significant. Life expectancy did not significantly differ across any two-year periods from 2015 through 2019. Life expectancy in 2020, however, was significantly lower than in all other years recorded in the study period (Figure 2). A substantial decrease in life expectancy—4.12 years (95% CI 2.06-6.57)—was observed between 2019 and 2020.

## Discussion

The impact of the COVID-19 pandemic on mortality in prison populations has not yet been well studied. To our knowledge, our study is the first to analyze how all-cause mortality rates, and thus life expectancy, changed in incarcerated populations in 2020, as well as the relationship between excess deaths and reported COVID-19 deaths. While the study population is limited to people confined in Florida state prisons, FLDOC has not reported exceptionally high rates of COVID-19-related infection or death as compared to other state prison systems.^21,22^ Similarly, Florida prisons’ population structure, in terms of age and sex distribution, is typical of other state prisons.^17^ As such, we anticipate that other prison systems are likely experiencing mortality patterns similar to those found in this study. At a minimum, similar investigation into other state prison systems is warranted.

Despite a population decline of more than 10,000 people between January and December 2020, FLDOC recorded more deaths among the prison population in 2020--590 individuals in total—than in any of the previous five years. During the 10 months of the COVID-19 pandemic in 2020, mortality rates were not only frequently much higher than would have been expected given previous years’ mortality trends, but the estimated excess mortality rate (∼42%) was also much greater than that estimated in the Florida population more generally (∼15%)^8^ or in the overall U.S. population (∼20%).^6^

To better contextualize the observed excess mortality in 2020, we can turn to life expectancy measures. Transforming yearly mortality rates to period life expectancy allows us not only to summarize the age-specific changes in mortality experienced by Florida state prisoners by way of life years lost, but also to compare those losses to other populations with different age structures.^23^ For example, in a recent publication by the CDC, it was estimated that in the U.S. population more generally, life expectancy declined by 1.0 years between January and June 2020, with a decline of 1.2 for males and .9 for females.^24^ Effects of this scale are historic and catastrophic. Yet our study found considerably greater declines in life expectancy, of 4.12 years, in the Florida state prison population. This again highlights the inordinately harmful impact of COVID-19 on the health of Florida’s prison population.

The staggering rate of excess deaths in FLDOC institutions raises the obvious question of what role the 191 COVID-19 deaths played in contributing to this excess. In the absence of COVID-19 infection, how many deaths may have been avoided? To quantify this value, detailed cause of death data across all years of the analysis would be needed to account for potential competing risks which may occur. Because cause of death data is unavailable to us, we are unable to assess the exact effect COVID-19 had on total mortality rates. Nevertheless, we found that for months with estimated high amounts of excess deaths, there was also a high reported number of COVID-19 related deaths (Figure 1). In addition, we know from previous studies that rates of COVID-19 infection have been substantially higher among the prison population than among the general population.^1^ Given these findings, it seems extremely likely that excess deaths would have been prevented had infection rates not been so widespread or had these individuals not been in a prison setting where the risk of COVID-19 infection was so high.

Several other factors should be considered as a possible explanation for the substantial increase in the Florida state prison mortality rate between previous years and 2020. First, while not specifically documented among the Florida state prison population in 2020, it is likely that, as in other prison populations, there is a disproportionately high prevalence of cardiovascular and respiratory diseases among those held in FLDOC facilities. Previous studies have documented that these conditions may increase risk of mortality for individuals with an active COVID-19 infection,^13–15^ and the greater overall increase in mortality in the prison population may be due in part to the higher prevalence of these pre-existing conditions.

Second, some of the change in the crude mortality rate can likely be attributed to a prison population that was substantially older in 2020 than in previous years. The decline in the size of the Florida prison population has largely been due to a drop in intakes rather than increased releases.^25^ As people coming into prisons tend to be significantly younger, it is not surprising that a drop in intake led to a decline in the number of younger prisoners, while older populations either remained relatively stable or slightly increased in number (Table 2). Given COVID-19’s high mortality rate for older individuals,^26,27^ this shift alone will have increased overall mortality risk in the prisons. This, however, only offers a partial explanation, as it only explains changes in the crude mortality rate and not the age specific mortality rate or life expectancy changes which account for changes in the age structure of the population.

### Limitations

Our analysis has several limitations. First, COVID-19 deaths were attributed to the month in which the deaths were reported and not necessarily to the month in which the individuals died. As such, it is likely that, in some instances, we have attributed COVID-19-related deaths to later months than the months in which they actually occurred. Second, because causes of death were not included in death data provided by FLDOC, we were unable to attribute age and sex to reported COVID-19 deaths. As a result, we cannot specifically attribute changes in age-specific mortality rates to COVID-19. Nevertheless, the age-specific increases in overall mortality rates are in line with changes in mortality rates due to COVID-19 found in other settings.^26,27^ Lastly, mortality events among the prison population may not be observed or recorded if an individual who is terminally ill is granted parole or compassionate release. If the pandemic occasioned a substantial year-to-year change in the number of individuals being released through either of these channels, our analysis might fail to accurately capture changes in the mortality risks to the Florida prison population. To date, however, there is no evidence of such a change.^28^

### Conclusion

To protect vulnerable incarcerated populations—including those held in FLDOC facilities—from COVID-19 and future viral pandemics, we must identify effective strategies for preventing infections in custody, for containing transmission, and providing adequate medical treatment for those with symptoms when outbreaks occur. While prison populations and density have declined in 2020, this study demonstrates that, in Florida at least, these declines have been insufficient to avert catastrophic impacts on mortality rates and life expectancy. Our findings thus offer evidence in support of calls from other experts for vaccine prioritization for the incarcerated and for strategic decarceration.^28–30^ This study demonstrates that people in prison have faced a substantially elevated total mortality risk during the COVID-19 pandemic, in addition to the elevated risk for COVID-19 infection found in previous studies.^1^ We urge officials in Florida and other states to evaluate interventions to reduce the risks associated with COVID-19 and to take steps to minimize the observed discrepancies in negative health outcomes between the general public and the incarcerated.

## Data Availability

All data concerning Florida state prison populations is made available by FLDOC. Historical records of COVID-19 death data for the Florida prison population is publicly available at https://github.com/uclalawcovid19behindbars/data.

https://github.com/uclalawcovid19behindbars/data

## Author contributions

SD, AL, and VR conceived the study. ME, VR, NM, HJ, and ET collected the data. NM, ET, and VR designed the study. NM conducted data analysis. NM drafted the manuscript with contributions from SD, AL, VR, ET, ME, HJ. SD and AL revised the manuscript. All authors reviewed the manuscript and agreed to be responsible for all aspects of the work.

## Acknowledgments

We would like to thank Sara Curran and Ben Nyblade for their insight into the analysis and to remember all those in the Florida state prison system who lost their lives to COVID-19.

## Appendix

Model formulation for excess mortality model.

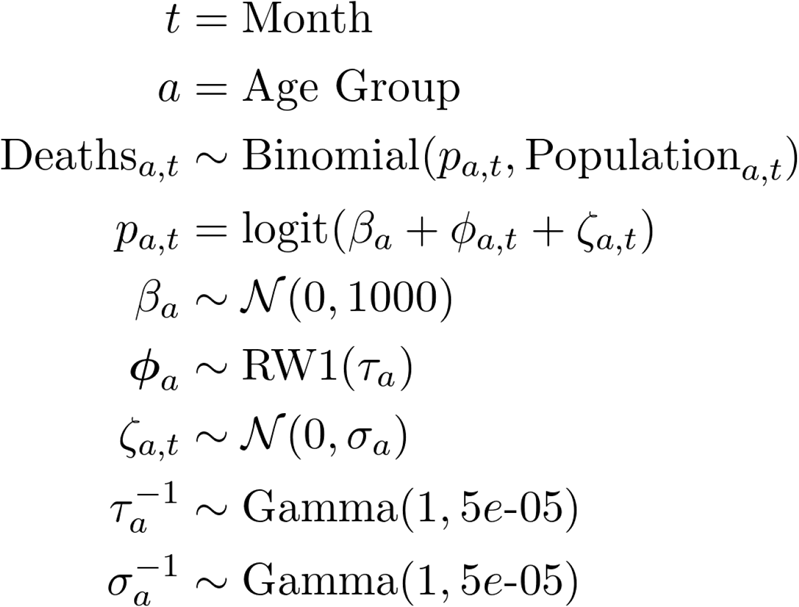

